# Exempting low-risk health research from ethics reviews to better serve the interests of the patients and public: a qualitative analysis of survey responses

**DOI:** 10.1101/2021.02.01.21250961

**Authors:** Anna Mae Scott, Iain Chalmers, Adrian Barnett, Alexandre Stephens, Simon E. Kolstoe, Justin Clark, Richard Matthews, Paul Glasziou

## Abstract

**Background:** We conducted a survey to identify what types of health research could be exempted from research ethics reviews in Australia.

**Methods:** We surveyed active Australian health researchers and members of Human Research Ethics Committees (HREC). We presented the respondents with eight hypothetical research scenarios, involving: N of 1 trials, no treatment studies, linked data sets, surplus samples, audits, surveys, interviews with patients, and professional opinion. We asked whether these scenarios should or should not be exempt from ethics review, and to provide (optional) explanations. We analysed the reasons thematically, to identify Top 3 reasons underlying the decisions.

**Results:** Most frequent reasons for requiring ethics reviews, included: the need for independent oversight, privacy/confidentiality issues, review of scientific rigour, and publishing considerations. Most frequent reasons for exempting scenarios from reviews, included: level of risk, study design, privacy/confidentiality issues, and standard clinical practice. Four research scenarios listed the same Top 3 reasons for requiring ethics reviews: need for independent oversight, review of scientific rigour, privacy/confidentiality. Reasons for exempting were less uniform, but low risk was a Top 3 reason for 7 scenarios, and study design for 4 scenarios. Privacy/confidentiality was given as a Top 3 reason for *both* requiring and exempting from ethics the same two scenarios.

**Conclusions:** The most frequently offered reasons in support of requiring ethics reviews for research scenarios are more uniform than those for exempting them. However, considerable disagreement exists about when the risks of research are so minimal that the exemption is appropriate.

## Introduction

Health research involving human participants generally requires review by a research ethics committee, to ensure that the participants’ rights and well-being are protected, and that the benefits of the research outweigh its harms. In Australia, this review is provided by human research ethics committees (HRECs), which assess the proposed research for compliance both with the applicable laws and the *National Statement on Ethical Conduct in Human Research*.(1)

Research ethics reviews conducted by HRECs can be likened to an intervention. Like other interventions – surgical procedures, pharmaceuticals, and so forth – they can not only benefit the patients and the public, but also come with drawbacks. Reported drawbacks of research ethics reviews include: impeding legitimate research through protracted time to decision, imposition of considerable costs and burdens on researchers and funders, a level of scrutiny that is disproportionate to the risks, lack of transparency around decision-making, roulette-like nature of decision-making, duplication of paperwork, committees’ being ill-equipped to review specific types of research, confusion around the roles of ethics committees, and scope creep.(2-9)

The impact of these issues in some cases will harm patients and the public by delaying access to effective therapies.(8) This has led some to conclude that in some cases, ethics review may do more harm than good.(10, 11)

Like harms from medical interventions, harms from the research ethics review processes can be identified through evaluative studies, so their impacts can be minimised or eliminated, and improvements can be made.(5, 8)

We report the results of a survey exploring the perceptions of Australian researchers and members of Australian research ethics committees on how the Australian research ethics system might better serve the interests of patients. Specifically, we wished to identify which types of research might be exempted from ethics review because the risks theoretically associated with them are minimal.

## Methods

We conducted an online survey of human health and medical researchers and members of HRECs working in Australia. Full methods are reported elsewhere.(12) Briefly, we identified active Australian researchers by searching PubMed for publications from 1 April to 30 June 2019, whose corresponding author’s affiliation included the words “Australia” OR “Australian.” This found 18,271 publications, which were exported to EndNote. The author email address field in EndNote was then searched for the term “.au” limiting the results to 4,956 references. From this pool, a sample of 900 names of researchers was randomly drawn.

HRECs were identified from the list of HRECs (n=208) maintained by the National Health and Medical Research Council, which contained contact information for many of the HRECs. Where contacts were unavailable, we used web searches. We identified contact information for 175 HRECs.

The survey presented 8 hypothetical research scenarios and respondents were asked whether each scenario should be exempt or not exempt from ethics review in Australia, and optionally, to provide reasons for their decisions. Abbreviated scenario descriptions are in Box 1 (for full descriptions, see Appendix 1). Because answering 8 scenarios was considered burdensome, we generated four versions of the survey, each containing 4 of the 8 scenarios. Researchers and HREC committees were randomly allocated to one of the four survey versions. The survey was conducted between 10 September and 1 November 2019.

### Box 1: Abbreviated* descriptions of the surveyed scenarios

**Scenario 1: N-of-1 studies in clinical practice (‘N of 1’ scenario):**

A GP randomises each of her patients taking statins, who have symptoms that may be side effects to 2 weeks of statin and 2 weeks of placebo, without disclosing the order to the patients. The patients record their symptoms during a 4-week period. The GP wishes to share her findings in a publication.

**Scenario 2: No treatment/no behavioural rules imposed (‘No treatment’ scenario)**

Participants in a study are not required to follow any behavioural rules that are not already practiced in routine care, nor receive any treatment that is outside routine clinical use. Study can be of any design (including a randomised controlled trial or observational study).

**Scenario 3: Linked data sets (‘Linked data sets’ scenario)**

Administrative data sets (such as electronic health records) are linked but deidentified. Original data sets were previously assessed for privacy, feasibility, etc. Individual data cannot be seen. Data are held in a secure facility and cannot be downloaded. Analyses can be performed but not downloaded.

**Scenario 4: Surplus samples or tissues during routine collection in clinical practice (‘Surplus samples’ scenario)**

Surplus (extra) tissue or samples (e.g. blood) obtained during routine procedures, not required for diagnosis or testing, being used for research purposes.

**Scenario 5: Quality assurance or audit project (‘QA/audit’ scenario)**

Audits to assess whether local care standards or practices are compliant with national standards or guidelines. Findings will be used for improvement and may also be published.

**Scenario 6: Survey/questionnaire of patients, lay persons or carer (non-professional) (‘Patient survey’ scenario)**

A survey involving patients, lay persons, or care providers, which does not include sensitive questions (e.g. mental illness, HIV status, etc.)

**Scenario 7: Interview with patients, lay persons or carers (non-professional) (‘Interview with patients’ scenario)**

An individual or group interview, where consent was obtained. Interview does not include sensitive questions (e.g. mental illness, HIV status, etc.)

**Scenario 8: Professional staff providing opinion/views in their area of expertise (‘Professional staff views’ scenario)**

Staff (e.g. researchers, hospital employees) are asked to volunteer their professional views. Information is collected via an interview or a survey.

*for a full description of the questions and scenarios surveyed, see Appendix 1.

Quantitative results from the survey have been reported elsewhere.(12) We report here the results of an analysis of the reasons offered in support of the respondents’ decisions to either exempt or not exempt the scenarios from ethics review. Each response was analysed thematically, using the inductive approach.(13) Two authors coded each comment to a theme; a third compared the two sets of codes, and rephrased or amalgamated them for consistency. The perspective is that of health and medical researchers. We corrected minor typographical errors in the quotes but otherwise made no changes.

We report here the Top 3 reasons offered by the respondents in support of their decisions to either require or exempt from ethics review, each of the eight scenarios. Where the top fourth most common reason was close to the top third most common reason in terms of the number of comments, it is reported here. Where there were several options for the top third most common reason for exempting or requiring ethics review for a scenario, and each had fewer than 5 comments, we did not report the top third reason.

[Blinded for peer review] Human Research Ethics Committee approved the project (32214912).

## Results

We successfully contacted 817 researchers and 175 HRECS (valid email addresses that did not bounce or auto-reply as no longer in service). We received 514 responses in total: 172 from researchers (21% response rate), and 342 from HREC members (estimated maximum response rate of 24%).

The number of responses (selections of either the ‘this scenario should require ethics review’ or the ‘this scenario should not require ethics review’ option) for the eight surveyed scenarios ranged from 189 to 250. The number of comments explaining the response or decision for each scenarios ranged from 119 to 181. (Table 1)

**Table 1:** Overview of the results

|  | Scenario 1: N-of-1 studies in clinical practice | Scenario 2: No treatment/no behavioural rules imposed | Scenario 3: Linked data sets ('big data' population health research) | Scenario 4: Surplus samples or tissues during routine collection in clinical practice | Scenario 5: Quality assurance or audit project (had 3 examples) | Scenario 6: Survey/questionnaire of patients, lay persons or carer (non-professional) (had 2 examples) | Scenario 7: Interview with patients, lay persons or carers (non-professional) (had 2 examples) | Scenario 8: Professional staff providing opinion/views in their area of expertise (had 3 examples) |
| --- | --- | --- | --- | --- | --- | --- | --- | --- |
| **Scenario abbreviation** | N of 1 | No treatment | Linked data sets | Surplus samples | QA/audit | Patient survey | Interview with patients | Professional staff views |
| **No. of responses** | 213 | 250 | 192 | 232 | 190, 189, 190 | 220, 221 | 195, 195 | 226, 227, 228 |
| **No. of comments** | 162 | 181 | 119 | 163 | 129 | 148 | 141 | 144 |
| **% of responses requiring ethics review** | 76% | 50% | 34% | 82% | 31%, 34%, 54%* | 41%, 71%* | 68%, 76%* | 16%, 15%, 60%* |
| **Top 3\*\* reasons offered in support of the decision to require review** | 1. Independent oversight<br>2. Review of scientific rigour<br>3. Publishing<br>4. Privacy / confidentiality | 1. Independent oversight<br>2. Review of scientific rigour<br>3. Privacy / confidentiality | 1. Independent oversight<br>2. Privacy / confidentiality<br>3. Review of scientific rigour | 1. Consent<br>2. Independent oversight<br>3. Review of scientific rigour | 1. Direct observation / interaction with patients<br>2. Publishing<br>3. Independent oversight | 1. Privacy / confidentiality<br>2. Independent oversight<br>3. Review of scientific rigour | 1. Independent oversight<br>2. Privacy/ confidentiality<br>3. Review but other than an HREC review | 1. Privacy/ confidentiality<br>2. Independent oversight<br>3. Publishing |
| **% of responses exempting from ethics review** | 24% | 50% | 66% | 18% | 69%, 66%, 46%* | 59%, 29%* | 32%, 24%* | 84%, 85%, 40%* |
| **Top 3\*\* reasons offered in support of the decision to exempt from review** | 1. Standard clinical practice<br>2. Level of risk | 1. Standard clinical practice<br>2. Study design<br>3. Exemption or minimal review | 1. Level of risk<br>2. Ethical considerations previously addressed<br>3. Exemption or minimal review | 1. Consent<br>2. Makes good use of waste<br>3. Level of risk | 1. Study design<br>2. Level of risk<br>3. Privacy / confidentiality<br>4. Standard clinical practice | 1. Level of risk<br>2. Study design<br>3= Privacy / confidentiality<br>3= Content of the questionnaire | 1. Level of risk<br>2. Consent | 1. Level of risk<br>2. Study design<br>3. Privacy/ confidentiality |

\*These scenarios had two or three examples – each was rated separately (see Appendix 1 for full descriptions). \*\*Where the fourth most common reason was close (in number of respondents citing that reason) to the third reason, it is reported here (e.g. N of 1 studies, QA/audit). Where there were several options for the third Top 3 reason, and each was cited by fewer than 5 comments, we did not report the third Top 3 reason (e.g. N of 1 studies, Interviews with patients scenarios).

## Most common reasons for requiring ethics reviews for the hypothetical research scenarios

### Independent oversight (8 of 8 scenarios)

The need for independent oversight by an HREC was cited as a Top 3 reason for the decision to require ethics reviews in all 8 of the surveyed scenarios.

#### Risk to participants

Many justified the decision to require HREC oversight for a scenario, because it involved a possible risk to participants. A respondent considering the N of 1 scenario stated *“Risk to participants requires a framework for oversight”;* another, discussing the QA/audit scenario stated that *“HREC review should ensure that foreseeable risks for participants are considered, identified and managed prior to commencement of the research*.” Considering the Patient Survey scenario, a respondent observed that *“Ethics can be used as a gateway and an opportunity for questions to be raised to ensure the safety of the patient/lay person/carer.”* In a similar vein, a respondent pointed out that the Professional Staff Views scenario is

> *asking staff about their work environment, interactions with peers, personal practices etc - all of which has potential to be unsettling or uncomfortable to the participant, so there is some risk and I think it should be reviewed by an ethics committee.*

#### Research rather than clinical practice

For some respondents, the independent oversight was required because the scenario described ‘research’ rather than clinical practice or treatment. One respondent observed that the N of 1 scenario *“is research, and requires independent review,”* and another pointed out that *“Research should be reviewed. It is not the same as treatment”* when clarifying their decision to require ethics review for the No Treatment scenario.

#### Reassurance to participants

Some respondents stated that having the independent oversight of an HREC is reassuring to the participants or the general public. A respondent stated that an ethics review for the Linked Data Scenario *“reassures patients whose data is used that an independent body had oversight and considered [t]he risks to them.”* Considering the Professional Staff Views scenario, a respondent observed stated that *“Staff may be more likely to participate if there has been an independent review.”* A respondent who required ethics reviews for the Surplus Samples scenario pointed out that *“Ethical review ensures public trust.”*

#### Concerns about researcher conduct

Several respondents required independent oversight provided by HRECs because of concerns about researcher conduct. In context of the Surplus Samples scenario, one respondent noted that such oversight *“keeps researchers honest. I would hate to think there was a potential for collecting extra blood or tissue with no accountability.”* Regarding the same scenario, another noted that ethics review *“is to protect participants from rogue/unethical research teams.”* More generally, a respondent on the Interview with Patients scenario observed that:

> *It is unrealistic to expect even good researchers of good character to be ethically infallible-oversight provides a helpful external perspective to maintain high standards of ethical care.*

### Privacy / confidentiality (6 of 8 scenarios)

Privacy and confidentiality issues were cited as a Top 3 reason for requiring ethics reviews for 6 of the 8 scenarios: N of 1, No treatment, Linked data sets, Patient Survey, Interview with Patients, and Professional Staff Views.

#### Potential to compromise privacy

Privacy concerns were raised about the N of 1 Studies scenario because *“The proposed research potentially compromises the privacy of patients and needs to be assessed by HREC to weigh risk to patients against public benefit.”*

#### Identifiability issues

Concerns about privacy and confidentiality were often framed by respondents as an identifiability issue. For example, worries were raised about the identifiability of the medical records in the N of 1 Studies scenario, which for some respondents raised *“Questions about whether medical records are identifiable”* which in turn were cited as a reason for requiring ethics review for those types of studies.

Identifiability concerns recurred in comments pertaining to the No Treatment scenario; respondents pointed out that the clinical information collected *“may include identifiable information.”* Similar comments were made about the Linked Data Sets scenario, as *“it may still be possible to identify individuals.”* Similar worries were raised in the Professional Staff Views scenario, as the staff:

> *need to be fully informed of the risks and guaranteed by the researchers that they will not be identified in any way and the monitoring role of the ethics committee approval process will provide some kind of confidence that this will happen.*

As a more general rule, in research scenarios where identifiability was a possibility, that was considered sufficient to require ethics review. For example, in the Interview with Patients scenario, a respondent stated that *“If person’s identity is a known factor (i.e. not deidentified) then ethics should be required.”*

#### Deidentification

Conversely, the possibility of deidentification was also offered as a basis for the decision to require ethics reviews. Commenting on the Linked Data Sets scenario, a respondent pointed out that:

> *Deidentification of data is a problematic concept, especially as the ability to cross link ‘deidentified’ datasets increases. Potential for reidentification is relevant.*

Similar concerns were echoed by respondents requiring ethics review for the Patient Survey scenario:

> *It is unrealistic in today’s world to believe that so-called ‘anonymous’ surveys cannot be reidentified by those with adequate skills.*

### Review of scientific rigour (5 of 8 scenarios)

The need to review scientific rigour was cited as a Top 3 reason for requiring ethics review in five scenarios: N of 1 studies, No treatment, Linked data sets, Surplus samples, and Patient Survey.

#### Review of methodology

Explaining their decision to require a review of the N of 1 scenario, a respondent said that “There absolutely has to be a review of the methodology, scientific merit, and ethical considerations of those patients” and another noted that there is a *“Need [for] peer review especially re: design, statistics to ensure adequate power to make conclusions.”* More generally, several respondents noted that the ethics review should be done *“To ensure the resultant outcome is scientifically useful.”*

A respondent who commented on the No Treatment scenario felt that:

> *the HREC review also considers the methodology, whether the study has sufficient statistical power to detect an effect etc. Without a robust statistical and scientific review the results may be useless, the resources consumed by the study wasted and the participants time and energy wasted for no potential for benefit making the study unethical.*

Along similar lines, a respondent required ethics review for the Linked Data Sets scenario as that scenario *“Requires evaluation of scientific merit and intended use [of the data].”* In context of the Surplus Samples scenario, another respondent specified that the following scientific and methodological issues would require consideration by the ethics committee: *“Circumstances of collection, storage, security, consent and disposal of samples.”* Respondents who required that an HREC review the Patient Survey scenario pointed out that *“an HREC can help determine if proper care was taken in designing the study.”*

#### Distrust in researchers’ methodological competence

Respondents sometimes required ethics review of the scientific rigour because they lacked trust in researchers’ methodological competence. One respondent justified their decision to require ethics review of the N of 1 scenario on the grounds that a *“GP [General Practitioner] may not be the best informed study designer”* – and another justified their decision to require an ethics review of the Patient Survey (and other research, more generally) by noting that *“in all these scenarios you are presuming a competent researcher.”*

### Publishing requirement (3 of 8 scenarios)

Publishing considerations were a Top 3 reason for requiring ethics reviews in three scenarios: N of 1, QA/audit, and Professional Staff Views.

#### Desire to publish

The desire to publish the findings was cited as a reason for requiring ethics review for the N of 1 scenario by a respondent who said that:

> *“The criteria that pushes this over a threshold for ethics review is the desire to publish and disseminate the research further. In this case, the methods/protocol should also be reviewed. Journals will often require evidence of ethics committee review.”*

Another respondent – who required ethics review for the Professional Staff Views scenario – stated that it: *“All depends on whether the researchers want to publish the outcomes.”* Commenting on the same scenario, another person pointed out that:

> *The presumption is that Ethics approval is needed when the results will be published or otherwise used for institutional purposes.*

#### Journal requirement

The completion of ethics review as a pre-condition for publication was sometimes attributed to a journal requirement. For example, one respondent who required ethics review for the QA/audit scenario asserted that *‘journals demand ethics approval,”* and another asserted that *“Studies aimed at publication will almost always need ethical review to enable publication.”* Considering the N of 1 scenario, a respondent similarly noted that the doctor in the described scenario *“may not be able to publish results without ethics approval and this would be a pity as I believe N of 1 has great potential to be informative.”*

#### Published project is research, which requires ethics review

At times, the reasoning to require ethics reviews for scenarios seemed to be underpinned by the following argument: if a project is being published, then that project is a ‘research project’ – and if a project is a ‘research project,’ then it requires ethics review. This is suggested, for example, by the following comment (about the N of 1 scenario):

> *I think that the desire to publish is (possibly) the key marker of what constitutes research. Researcher[s] usually need to (or feel the need to) add numerous steps, tests or other inconveniences to participants to get the addition angle that makes something routine into publishable material.*

This was stated more explicitly in the following comment explaining the decision to require ethics reviews for the QA/audit Scenario:

> *If it [the findings of the audit] is being used for research publication purposes it is research and therefore requires ethics approval.*

And similarly,

> *If the findings are published, the data is therefore human research data and that activity should require review.*

## Most common reasons for exempting the hypothetical research scenarios from ethics reviews

### Level of risk (7 of 8 scenarios)

The level of risk was offered as a Top 3 reason for exempting from ethics review in seven of the eight scenarios. The sole scenario for which the level of risk was not among Top 3 most commonly cited reasons for exemption was the No Treatment scenario (the level of risk was cited as a reason for exempting this scenario, although it was not a Top 3 reason – see Table 1).

#### Low or negligible risk

Some of the respondents referred to the low or negligible level of risk in the described scenario, as a reason for the decision to exempt that scenario from ethics review. For example, the N of 1 scenario was said to pose *“Low risk to participants,”* the Linked Data scenario was considered to have an *“inconceivable risk of potential harm”*, and the Patient Survey scenario was regarded as having *“no potential for harm to the participants.”* Respondents also noted, regarding the QA/audit scenario, that *“no risk to participants is introduced”* and similarly, considered that in the Surplus Tissues scenario, *“there is no additional procedure or risk for participants.”* The Professional Staff Views scenario was said to *pose “No tangible risk to human health. This sort of things goes on all the time by non-researchers who never seek ethics approval anyway.”*

#### Low or negligible risk and the potential for benefit

Several respondents coupled their reasoning about low risk inherent in the scenario, with the additional potential for benefit. Considering the QA/audit scenario, a respondent pointed out that it has *“No risk to patients; potential for future patient benefits,” and* regarding the Patient Survey scenario, another observed *that “Risks are negligible; potential for benefit/s.”* Respondent considering the Interviews with Patients scenario, noted the potential benefit to the interviewees from discussing their experiences:

> *“Interviews are not risky when consent is given. People can choose what they want to talk about. You are only asking about personal experiences - respondents are already aware of their own personal experiences and usually discussing these is only helpful, even when it is sensitive.”*

Others identified the benefits from decreased research costs. In context of the Patient Survey scenario, a respondent stated that:

> *The risk to respondents is extremely low. The general populace is now very familiar with completing surveys so is unlikely to cause any undue anxiety for the respondents. Removing the requirement for an ethics review decreases the cost of conducting research*.

#### Ethics reviews are a barrier

Many respondents identified the requirement for ethics reviews in these scenarios as a barrier to conducting the work. Considering the Linked Data Sets scenario, a respondent noted that:

> *Secondary analysis of deidentified data is one clear case where ethical approval should not be required. There is no risk to participants and ethical approval is an unnecessary barrier to conducting this research.*

Another made a similar observation about Interviews with Professional Staff, pointing out that requiring ethics review for this type of project turns individuals off conducting such projects:

> *ethics for this reduces this [type of] work being done as researchers[are] turned off doing this due to ridiculous ethics requirements*

Removing the ethics review requirement for Interviews with Patients scenario was also considered to be beneficial in terms of decreased burden to the HREC’s workload, as the scenario was considered:

> *Very low risk. We see a lot of applications like this and I think that the administrative burden is not justified by the level of risk.*

#### More general rule

A more general rule around ethics reviews and level of risk was suggested by a respondent to the Patient Survey scenario:

> *any approach that is not invasive nor provides risk, can in my opinion be exempted. Assuming that a researcher, as part of an organisation, operates in an ethical way (which is part of a professional code)*

### Study design (4 of 8 scenarios)

Study design was a Top 3 reason for exempting the following scenarios from ethics reviews: No treatment, QA/audit, Patient Survey, and Professional Staff Views.

#### Perceived study design: quality improvement or audit

Although the No treatment scenario explicitly stipulated that it could involve any study design (including a randomised or observational design), many respondents exempted it on the grounds of perceived study design. For example, one respondent considered that:

> *This study is best viewed in terms of a clinical audit comparing current practices. As long as patients are randomly allocated to the 2 groups I don’t think Ethics review is required.*

Others considered the scenario to be *“an audit”* or a *“Quality improvement program,”* and offered this as a reason for exempting the scenario from ethics review.

Those exempting the QA/audit scenario on the grounds of study design, noted that “these sorts of audits should not require ethics review,” or pointed out that *“quality improvement projects are subject to institutional governance oversight - which is sufficient.”*

Respondents to the Patient Survey scenario similarly pointed out that *“Surveys are used regularly as part of quality initiatives”* in support for exempting the scenario.

Several respondents considered the Professional Staff Views scenario to describe a quality improvement programme, exempting it from ethics reviews on that basis. One stated that *“These studies are an essential part of quality improvement and enhance clinical practice,”* whilst another commented that *“These are quality assurance projects enhancing research methods and clinical practice and have no ethical impact on participants.”* Similarly, others pointed out that *“they are predominantly QA processes” and “These are either informing research priorities (as in [example] A) or evaluating a program or initiative which falls under improvement [examples B, C].”*

#### Ethics review as an obstacle

Requiring ethics review for these types of study designs was considered an obstacle by some respondents. One respondent pointed out that:

> *Critical audit such as this is primarily to improve standard of care, and obtaining ethics review may be an obstacle to those wanting to do such audit*

Another respondent commented along similar lines that:

> *These are quality improvement activities. Burdensome governance reduces the likelihood of this sort of process and outcomes evaluation proceeding. Positive change opportunities are reduced if audit/review/evaluation is made too cumbersome.*

### Privacy / confidentiality (3 of 8 scenarios)

Privacy and confidentiality considerations were a Top 3 reason for exempting three scenarios from ethics reviews: QA/audit, Patient Survey, and Professional Staff Views.

In support of the decision to exempt the QA/audit scenario, a respondent noted that *“none of these [examples within the scenario] are taking any personal information”* and another pointed out that *“identities of patients are protected.”* More generally, *“[p]rovided that the data is securely stored and de-identified, this is what the public expect of those in the medical fields to maintain standards.”*

Commenting on the Professional Staff Views scenario, a respondent stated that they *“Do not believe the actions will significantly compromise anyone”* and another justified the decision to exempt by drawing an analogy to everyday conduct:

> *No issues around consent, privacy or potential harm. These are not unlike conversations we would have in everyday life.*

Regarding the Patient Survey scenario, a respondent who exempted the scenario noted succinctly that the *“privacy of participants is protected” and another stated that “OK [to exempt] if individual responses not identifiable.”*

This scenario was considered manageable with respect to issues around privacy or confidentiality, and also to describe conduct that should be expected of staff:

> *Data can be anonymous and de-identified, as health care providers and professionals there should be an expectation to participate in this type o[f] research to improve healthcare delivery.*

### Standard clinical practice (3 of 8 scenarios)

Three scenarios were commonly exempted from a requirement to undergo ethics reviews on the grounds that they describe standard clinical practice: N of 1, No treatment, and QA/audit.

A respondent observed that the N of 1 scenario *“falls within standard clinical practice. The intention of the researcher (as presented) is clinical treatment. Note-taking is a good part of this.”* Another stated that:

> *This is part of necessary treatment/investigations for the patient. Delaying this investigation for ethics application is unnecessary and detrimental to the patients - this will probably take 6 months at least for Ethics review!*

In supporting the exemption for the No Treatment scenario, a respondent pointed out that:

> *Nothing is being done that would not be done anyway, except, basically, counting. I don’t see any danger here, especially if the hospital is just using supplies that it would be using anyway. There does not seem to be any risk to anyone that exceeds the risk the hospitalised patient faces simply from being in the hospital.*

Similar observations underpinned the decisions to exempt the QA/audit scenario from a requirement to undergo ethics reviews. These types of scenarios *“Do not involve alterations to patient trajectory or treatment options”* and *“All procedures are being done as part of standard care; identities of patients are protected no change to patient care and intent to improve local practice.”*

## Discussion

Exemption of scenarios from ethics review was most commonly justified by their low risk (a Top 3 reason in 7 of 8 scenarios). Other justifications were less uniform across scenarios, but included: study design (a Top 3 reason in 4 scenarios), privacy/confidentiality considerations (in 3 scenarios) and standard clinical practice (in 3 scenarios). Conversely, the reasons offered for requiring ethics reviews were more uniform – for example, the need for independent oversight was cited as a Top 3 reason for all 8 scenarios. Moreover, four of the scenarios (N of 1, No Treatment, Linked Data Sets, and Patient Survey) all list the same Top 3 reasons for requiring ethics reviews, albeit in differing order. Considerations of privacy/confidentiality were cited as a top reason for both exempting from and requiring ethics reviews for two scenarios: Patient Survey and Professional Staff Views (see Table 1, and Table A1, Appendix 3).

An unexpected finding was the pervasiveness of the view that ethics reviews should be conducted because HRECs provide independent oversight – a Top 3 reason for requiring ethics reviews for all 8 scenarios. This is particularly problematic for the N of 1 trials scenario, as due to co-morbidities and genetic makeup, the harms experienced by individual patients will differ from the harms identified in studies with larger groups (e.g. RCTs). Robustly conducted, randomised and placebo-controlled N of 1 trials, can resolve the question of whether it is that treatment that causes those harms in that particular patient. Requiring ethics reviews for N of 1 trials may therefore perversely incentivise the conduct of informal, scientifically inferior, poorly controlled trials – which do not require ethics reviews, and are therefore easier to set up and conduct – rather than robustly designed ones – which require ethics reviews and the associated expense in time and resources.(14)

Another unexpected finding was the frequently expressed view that in order to be published, a project must have obtained ethics approval. Several respondents justified their reasoning with the following argument: if a project is being published, then it is ‘research’ – and if a project is ‘research,’ then it requires ethics review. Although logically, it would follow from those premises that if a project is being published then that project requires ethics review, both premises are problematic. First, it is not the case that only research projects are published – plenty of other work is published, including audits, quality improvements, quality assessments, and so on. Second, it is not clear that ‘research’ ipso facto requires ethics reviews – systematic reviews almost always do not, and a wide range of what is uncontroversially considered to be ‘research’ (e.g. randomised controlled trials, work with biospecimens, surveys, interviews, etc.) is exempted from ethics reviews in various countries subject to meeting prespecified criteria.(1)

To the best of our knowledge, this is the first study to evaluate researchers’ and ethics committee members’ views on potential exemptions from ethics review in Australia of research currently exempted in other countries, although how these groups perceive the role and function of ethics reviews committees has previously been reported,(3) as have the experiences of researchers in social sciences and humanities with Australian human research ethics committee processes.(15) Our survey therefore contributes to the small but growing pool of evaluations of research ethics reviews – both in Australia and overseas – which, despite calls for their conduct(16, 17) remain relatively rare.(18) One of the reasons for this is that research ethics bodies are reluctant to participate in this reflective research. For example, in a recent study of how Institutional Review Boards function in the United States, only 40% of Boards agreed to participate.(19) This is consistent with our experience, as our maximal response rate from HREC members was approximately 24%. Other commonly identified difficulties with conducting this type of research is that there is much uncertainty about the best methods to do so(17, 18, 20) and absence of a ‘gold standard’ for evaluating research ethics review processes.(6) Despite these challenges, such studies are vital, as they help identify the shortcomings of the current approaches – and point towards opportunities to mitigate them.

Our study had some limitations. First, our survey respondents may have had stronger feelings about the *status quo* in Australia, or be more interested in ethics reviews, than non-respondents, which could result in a biased sample, and limit the generalisability of our conclusions. This was partly mitigated by the use of a random (rather than convenience) sampling for researchers, although randomising was not practical for the HREC respondents and instead we tried to contact all committees. Second, because the issues around research ethics processes in Australia are of most immediate impact on researchers and research ethics committee members, those were the groups we surveyed. However, it is the patients and the public who are ultimately impacted by the efficiencies or inefficiencies in the ethics review processes (e.g. via prompt or delayed access to assessment of therapies), and their views may not be in alignment with the views expressed by the respondents to our survey. Finally, we deliberately did not collect any demographic information, other than whether the respondent was a researcher, an HREC member, or both, as part of our survey. Although this precluded an assessment of whether our respondents were representative of the broader research and HREC communities, we did so to maximise respondent privacy and encourage candour.

One immediate outcome of this research has been a letter sent to HRECs, to address the misconception that journals always require research ethics approval. On 8 and 9 September 2020, two of the authors (AB, AMS) sent a letter to the 175 Australian HRECs originally surveyed (see Appendix 2), with a sampling of quotes from our respondents, and the clarification that medical journals often allow authors to explain why their work was exempt from ethics review, and publish such work. Fourteen HREC contacts responded to confirm the receipt of our letter, and advised us that they have disseminated the letter to their members or discussed it during a meeting. We have also provided the manuscript with the results of this survey to the National Health and Medical Research Council, whose *National Statement on Ethical Conduct in Human Research* describes the human research ethics principles and HREC review processes in Australia, and is currently being revised. Finally, as we recognise that the researchers and the HRECs are often siloed, we are currently in the early stages of planning a joint meeting of researchers and HRECs, to identify areas that both sides could agree on which priorities should be addressed and the appropriate means of doing so.

The range of opinions among researchers and members of research ethics committees in Australia provided in our survey makes it clear there is considerable disagreement about the circumstances in which any risks associated with proposed research are plausibly so minimal or non-existent that the studies should be exempt from ethics review. It is not clear how best to resolve this, although exempting these proposed studies from ethics review would have the benefit of freeing up resources for ethics review to focus on, for example, ensuring that the methods of proposed studies have been reliably assessed by research funders and sponsors before ethics review. This could mitigate some of the HREC workload, and additionally, address the concerns raised by many of the survey respondents, who cited the review of scientific rigour as a Top 3 reason in support of decisions to require ethics reviews.

## Conclusion

Innovation in research ethics review processes is challenging although not impossible. Randomised controlled trials are generally considered higher risk and undergo extensive ethics review prior to commencement, but the recent RECOVERY trial, which aims to identify treatments for hospitalised COVID patients, obtained ethics approval and enrolled first patients in 9 days.(21) Streamlining approvals for low risk research therefore should be possible. The status quo – of burdensome, complex, not fit-for-purpose system of ethics reviews – need not be the default. We *can and must* progress toward more evidence-based, fit-for-purpose, and patient- and public-oriented ethic review processes.

## Data Availability

Deidentified data will be available upon reasonable request from the corresponding author.

