## Appendices for "Exempting low-risk health research from ethics reviews to better serve the interests of the patients and public: a qualitative analysis of survey responses"

### Appendix 1 – Full scenario descriptions

**Scenario 1: N-of-1 studies as part of regular, clinical practice**

A patient recently started on a statin has complained to his general practitioner (GP) about muscle pains and fatigue, and says that he is considering stopping the drug. The GP wants to be confident that the statins are likely to be the cause of the patient’s symptoms, so – with the patient’s consent – the patient is allocated to 2 weeks on a placebo and 2 weeks on a statin, but with the ordering of the drug and placebo concealed. Altogether, 20 of the GP’s patients taking a statin and complaining about the unpleasant symptoms, consent to using this approach before deciding whether to stop the statins. The patients keep records of their symptoms throughout the duration of the administration of the drug or placebo. The GP finds that most patients experience unpleasant symptoms whilst taking statins, but the symptoms disappear whilst taking placebo. The GP wishes to use these findings to help her patients and share them more widely through publication.

**Scenario 2: No treatment/no behavioural rules imposed**

Study of any design (cohort, randomised control trial, other designs) that does not require participants to receive any treatment that is not already in routine use, and does not require participants to follow any behavioural rules that are not already applied in routine care.

For example, consider a study involving hospitalised patients. The hospital already uses two types of mattress, with some beds using mattress A, and other beds using mattress B. There is uncertainty about whether one of the mattresses is better at preventing bed sores than the other. To address this uncertainty, patients will be randomly allocated either to mattress A or to mattress B, and the incidence of bed sores will be compared in the two groups.

**Scenario 3: Linked data sets (‘big data’ population health research)**

A study involving the use of linked, de-identified administrative data sets (e.g. registries, administrative data, electronic health records).

For example, consider a study of the safety of the cardiovascular safety of non-steroidal anti-inflammatory drugs (NSAIDs e.g. Naprosyn, Nurofen). This study involved a de-identified (no names, full date of birth, postcode) data set of people over 75 years of age who used, or did not use, NSAIDs. The prescription records were linked to hospitalisation diagnoses and deaths (with causes) and the rates of fatal and non-fatal heart attacks were compared in users and non-users of the drugs.

The data centre holding the original data set has assessed the proposed project and analyses for privacy impact, feasibility of the study with the available data, and adequacy of data set creation and analysis plans, and has approved the project. The de-identified linked data were held in a secure facility. Individual level data could not be seen on screen. Data were located in a secure environment that allowed analysis but did NOT allow download of the data and did NOT allow download of the analyses. The analyses were checked by data-centre staff for privacy risk before release to the researchers.

**Scenario 4: Surplus samples or tissues during routine collection in clinical practice**

Research may use surplus (extra) tissue or samples obtained from people during routine medical procedures. For example, extra blood could be drawn at the time of medically required or routine sampling. Tissue not required for diagnosis or testing could be collected during routine clinical procedures requiring tissue sampling.

**Scenario 5: Quality assurance or audit project**

To promote good clinical care, audit projects are done to produce information about whether care standards comply with national standards and practice guidelines. Those conducting the audit will report the findings to inform planned improvements locally, and they may also disseminate their findings through publication in a journal.

Example A: An audit and feedback project to reduce inappropriate prescribing, e.g., a chart audit of sedative-hypnotics prescribed to older hospitalised patients.

Example B: Evaluation, pre- and post-simulation training, of door-to-needle time of patients with acute stroke, with the aim of improving treatment initiation.

Example C: Use of direct observation to examine nursing care for sick newborns, to identify missed elements of care, such as phototherapy sessions.

**Scenario 6: Survey/questionnaire of patients, lay persons or carer (non-professional)**

A survey or questionnaire of patients, lay persons, or care providers, which does NOT include questions about highly sensitive areas (e.g. suicide, mental illness, HIV status, recreational drug use), AND meets one the following criteria:

Example A: Identity of respondents cannot be readily identified (i.e. the survey or questionnaire is anonymous)

Example B: Identity of respondents is known only to the research team and is not disclosed outside of the research team

**Scenario 7: Interview with patients, lay persons or carers (non-professional)**

An interview (one-on-one or in a group, such as a focus group) of patients, lay persons, or care providers. Consent has been obtained from participants by the research team. The interview does NOT include questions about highly sensitive areas (e.g. suicide, mental illness, HIV status, recreational drug use), AND meets one of the following criteria:

Example A: identity of interviewed person(s) is known to the research team but not to the interviewer, and identity cannot be readily identified by the interviewer,

Example B: identity of interviewed person(s) is known to the research team including the interviewer, but is not disclosed to anyone else

**Scenario 8: Professional staff providing opinion/views in their area of expertise**

Professional staff (e.g. hospital staff, researchers) are asked – as volunteers – about their views on a topic in their areas of expertise or professional practice, through either a face to face interview or written/online survey. For example:

Example A: A survey of general practitioners to prioritise a list of systematic review topics for a Cochrane group.

Example B: A survey asking systematic reviewers to report on the strengths and weaknesses of systematic review software.

Example C: An interview of all physiotherapists in a hospital on barriers and facilitators they have experienced in implementing a specific therapy for patients.

### Appendix 2 – Letter disseminated to HRECs

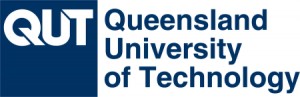

Professor Adrian G Barnett

**Queensland University of Technology**

09 September 2020

Publication requirement for ethical clearance

Dear Colleague,

In late 2019, we sent to you a request to disseminate a survey concerning your HREC members’ views on which low-risk research projects could be exempt from ethics review. We also sent this survey to Australian health researchers. Our survey focused on low-risk research, such as re-analyses of existing data, quality assurance, and online surveys.

We are writing to you for two reasons.

First, we would like to thank you very much for sharing our survey with your HREC members. We received over 500 responses, and many insightful comments. The survey results are now undergoing peer review with a journal, and have been published in a preprint^1^.

Second, an unexpected finding from the survey was that many researchers and ethics committee members expressed the view that if a project’s findings were to be published in a journal then ethics approval was required, even if the project was negligible or low risk.

This was evident from comments such as:

- “The criteria that pushes this over a threshold for ethics review is the desire to publish and disseminate the research further.”
- “Studies aimed at publication will almost always need ethical review to enable publication.”
- “Journals demand ethics approval.”

It is not the case that ethics approval is always required for publication. Medical journals do allow authors to explain why their project was exempt from ethics review, and regularly publish findings that did not require or receive ethical review.

We are writing in the hope that you will make your committee members and researchers aware of this anomaly. Exempting truly low-risk research from ethical review will be of benefit to both HREC members and researchers.

Yours sincerely,

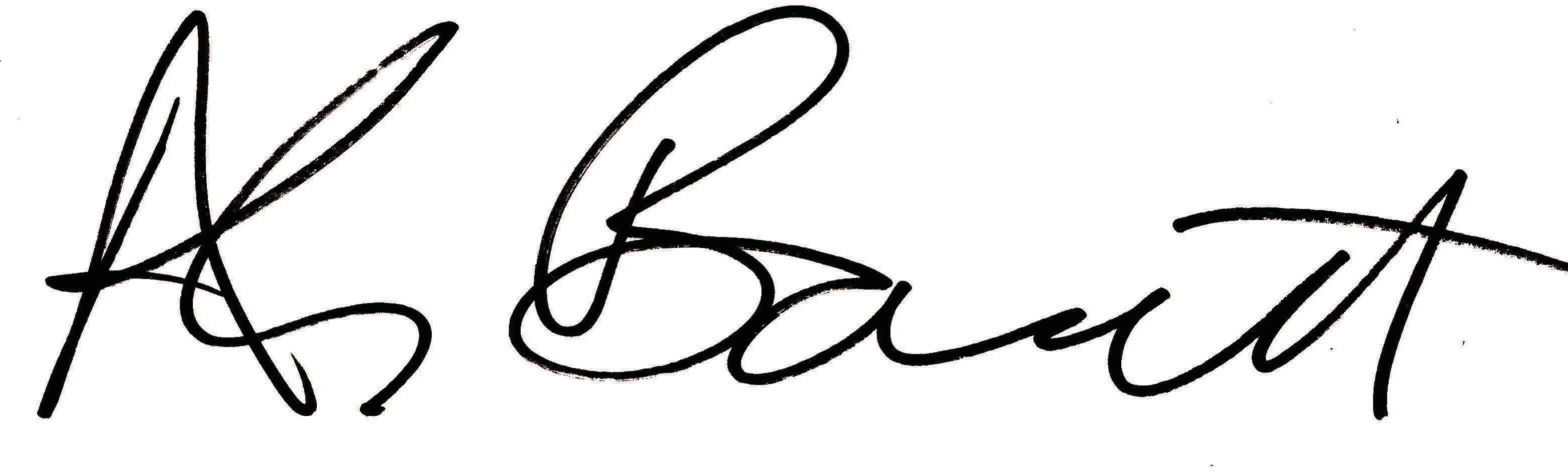

Adrian Barnett (QUT) & Anna Scott (Bond University)

**1 h**[**ttps://www.medrxiv.org/**](http://www.medrxiv.org/content/10.1101/2020.07.22.20159533v1.full.pdf)**conten**[**t/10.1101/2020.07.22.20159533v1.full.pdf**](http://www.medrxiv.org/content/10.1101/2020.07.22.20159533v1.full.pdf)

### Appendix 3 – Top 3 reasons for exempting from or requiring ethics reviews across all 8 hypothetical research scenarios

**Table A1. Top 3 reasons for exempting from or requiring ethics reviews across all 8 hypothetical research scenarios**

| **Number of scenarios for which it was a Top 3 Reason** | **Top 3 reasons for exempting research scenarios from ethics reviews** | **Top 3 reasons for requiring ethics reviews for research scenarios** |
| --- | --- | --- |
| 8 | ----------------- | Independent oversight |
| 7 | Level of risk | ----------------- |
| 6 | ----------------- | Privacy / confidentiality |
| 5 | ----------------- | Review of scientific rigour |
| 4 | Study design | ----------------- |
| 3 | Privacy / confidentiality  Standard clinical practice | Publishing |
| 2 | Exemption or minimal review  Consent | ----------------- |
| 1 | Ethical considerations previously addressed  Makes good use of waste  Content of the questionnaire | Review other than HREC review  Direct observation/interaction with patients |
